# A closer look at weight loss interventions in primary care: A systematic review and meta-analysis

**DOI:** 10.1101/2022.08.03.22278290

**Authors:** Leigh Perreault, E. Seth Kramer, Peter C. Smith, Darren Schmidt, Christos Argyropoulos

## Abstract

**Background:** Weight loss observed in clinical trials has failed to translate into real world clinical settings. The objective of the current review was to quantify patient weight loss using various approaches tested in primary care and to unveil relevant contextual factors that could improve patient weight loss on a long-term basis.

**Methods:** Data were compiled from Cochrane Central Register of Controlled Trials, MEDLINE, EMBASE, Scopus, and Web of Science from inception to October 30, 2022. Only randomized clinical trials conducted in people with overweight or obesity in a primary care setting where the intervention was administered by a primary care provider for at least 6 months were included. All investigators identified studies and independently abstracted data using COVIDENCE systematic review software. Quality assessment and risk of bias of individual studies was assessed in the context of the primary outcome using the template provided by COVIDENCE.

**Results:** The seven studies included 2,187 people with obesity who had weight-related comorbidities or risk factors. Substantial heterogeneity in the outcomes was observed, as well as bias toward lack of published studies showing no effect. The random effect model estimated a treatment effect for the aggregate efficacy of primary care interventions −3.54 kg (95% CI: - 5.61 kg to −1.47 kg). Interventions that included a medication component (alone or as part of a multipronged intervention) achieved a greater weight reduction by −2.94 kg (p<0.0001). In all interventions, efficacy declined with time (reduction in weight loss by 0.53 kg per six months, 95%CI: 0.04-1.0 kg, p-value=0.04).

**Discussion:** Weight loss interventions administered by a primary care provider can lead to modest weight loss. Weight loss is approximately doubled if anti-obesity medication is part of the treatment strategy. Nevertheless, weight loss is attenuated over time and underscores the need for long-term treatment.

## INTRODUCTION

Obesity continues to cast an enormous human and economic toll. As of 2018, the Centers for Disease Control and Prevention estimated that 73.6% of U.S. adults are considered overweight (e.g. body mass index (BMI) ≥ 25 kg/m^2^) and 42.4% have obesity (e.g. BMI ≥ 30 kg/m^2^)^1^, collectively costing the health care system approximately $1.7T annually. ^2^ Obesity is being increasingly recognized not only as a risk factor for disease but a disease unto itself. ^3^ Despite this fact, only ∼50% of people with a BMI of 50 kg/m^2^ (considered morbid or extreme obesity) have a diagnosis of obesity^4^ and <1% of people with any degree of overweight or obesity are offered anything other than lifestyle advice. ^5^ Given the paucity of certified “obesity expert” health care providers, this treatment gap can only be closed by systematically addressing the barriers to weight management in primary care.

Patient demand is high for weight management in primary care. A study by Sherson et al reported that most patients want to discuss weight loss with their physicians. ^6^ Specifically, patients value physician direction with their diet, physical activity and goal setting. ^7^ In addition, recent years have ushered in numerous and diverse options for weight management that extend signficantly beyond lifestyle advice. Medications for weight loss are being more extensively studied and demonstrating improved outcomes. ^8-11^ Bariatric surgery provides a possible option to address weight and reverse potentially life-threatening conditions such as heart disease and diabetes in both adolescents and adults. ^12-15^ Intensive behavioral therapy (IBT) for obesity is now a covered benefit under Medicare.^16^ Together, primary care practice transformation, reimbursement for obesity care and better therapies for weight management suggest new and pragmatic approaches can emerge.

Weight loss observed in clinical trials has failed to translate into real world clinical settings. ^5^ Most of these trials have tested one or more strategies for weight loss but not the context in which the strategies will ultimately be deployed. The objective of the current systematic review and meta-analysis is to determine weight loss in randomized clinical trials specifically conducted in a primary care setting where a primary care provider adminstered the intervention for at least six months. Our goal was to quantify patient weight loss using various approaches tested in primary care and to unveil relevant contextual factors that could improve patient weight loss on a long-term basis.

## METHODS

### PRISMA reporting guideline

This systematic review is reported according to the Preferred Reporting Items for Systematic Reviews and Meta-analyses (PRISMA) statement extension for network meta-analysis ^17^ and was conducted following an *a priori*–established protocol registered with PROSPERO (CRD42021242344).

### Selection criteria

Randomized clinical trials were included in this meta-analysis if they were conducted in a primary care setting with the primary care provider (physician, physician assistant, or nurse practitioner) administering the intervention for at least six months. Interventions administered exclusively by a medical assistant, health coach, dietician, or behavioral health provider were not included, even if they occurred in a primary care setting. Interventions referring patients elsewhere (e.g. to a commercial weight loss program) were also not included, even if the referral was placed by the primary care provider. The control condition may have been a placebo pill and/or some minimal amount of lifestyle intervention offered to both groups intended to resemble “usual care”. This analysis is limited studies to included adults age ≥18 years old with a BMI ≥25 kg/m^2^, with or without weight-associated comorbidities, that reported absolute weight at baseline and at the end of follow-up in the intervention and control groups.

### Search strategy

The search strategy was designed in consultation with an experienced medical librarian with input from study investigators using various databases from inception to October 30, 2022. COVIDENCE systematic review software (Veritas Health Innovation; Melbourne, Australia) was used for screening, full-text review, and data extraction (Figure 1). COVIDENCE is the primary tool used in Cochrane reviews accessing the Cochrane Central Register of Controlled Trials and databases supporting the Central Register such as MEDLINE, EMBASE, Scopus, and Web of Science. References were imported using the search terms: randomized clinical trial, obesity, overweight, weight loss, primary care, general practice, family practice, internal medicine, routine medical practice. Duplicate citations were removed. Studies advanced to full-text review as long as weight change was the primary outcome. Further exclusions were applied to ensure that only randomized controlled clinical trials conducted in a primary care setting with a primary care provider administering the intervention for ≥6 months were retained (Figure 1).

**Figure 1.**
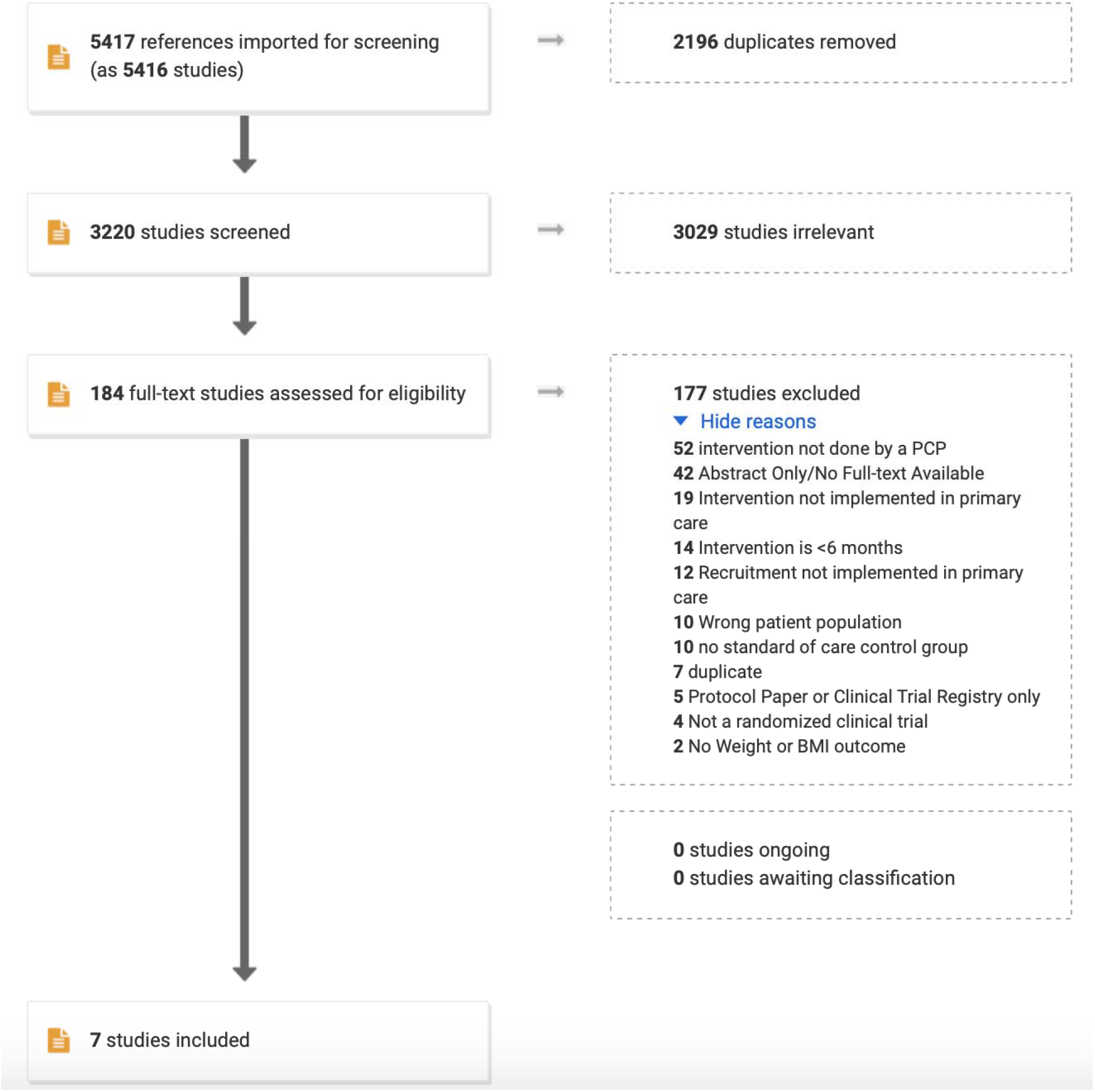
PRISMA diagram for data handling.

### Data abstraction and quality assessment

All authors participated independently in the screening and full-text review process described above. Discrepancies were resolved by consensus. Quality assessment and risk of bias of individual studies was assessed in the context of the primary outcome using the template provided by COVIDENCE. The template stratified responses into high, low or unsure for sequence generation (the method used to generate the allocation sequence should produce comparable groups), allocation concealment (the method used to conceal the allocation sequence should not have been foreseen in advance of, or during, enrollment), blinding of participants and personnel (neither participants or personnel had knowledge of which intervention a participant received), blinding of outcomes assessment (outcome assessors had no knowledge of which intervention a participant received), incomplete outcome data (data were complete with low attrition and exclusions from the analysis), selective reporting (low probability of selective outcome reporting), and other sources of bias (no concerns about bias not addressed in the other domains in the tool).

### Outcomes

All outcomes were assessed at 6 months of intervention vs. control, and again at 12, 18, or 24 months as long as the intervention vs. control conditions were ongoing according to the randomization. The primary outcome was weight change from baseline. Contextual factors of interest focused largely on strategy employed in the intervention group, but patient characteristics and control condition were also examined. When available, outcomes were abstracted using study-reported modified intention-to-treat analysis (i.e. patients exposed at least once to the intervention or control condition and had one post-randomization weight assessment).

### Statistical analysis

Publication bias was assessed using contour-enhanced funnel plots ^18^ to detect the area of statistical significance in which studies were missing, and the statistical significance of funnel plot asymmetry was analyzed with the random effects version of the Egger test. ^19^ To generate this plot multiple treatment effects from individual studies (change from baseline relative to control treatment at multiple time points and/or multiple interventions) were synthesized via random effects meta-analysis to provide an overall treatment effect per study. This approach was used to generate a summary forest plot from all studies considered. The fixed effects model assumes that the treatment effect is the same across all trials, while the random effects model assumes that the treatment effect varies between the trials. Statistical heterogeneity within and across trials synthesized was quantified by the I^2^ statistic and the between study variance(τ^2^), which was calculated by REstricted Maximum Likelihood (REML). The p-value of the Q test was used to test for the statistical significance of the observed heterogeneity. To explore the replicability of these findings we calculated the prediction interval ^20^ (i.e. the 95% confidence interval for the treatment effects in a future trial). A prediction interval that is narrow and overlapping with the confidence interval of the treatment effect, suggests that future studies are unlikely to change the conclusion we can draw from the currently available trials. Finally, a formal, multilevel meta-regression analysis was carried out to explore the hypothesis that lifestyle modifications will have a different effect than pharmacologic interventions. This hierarchical meta-regression was the primary means for synthesizing evidence for our analysis and accounts for the correlation of outcomes due to the common control group in interventions with multiple treatments assessed at multiple time-points during each study. The analyses were conducted in Microsoft R open v4.0.2 packages meta v4.13-0 (funnel plot/forest plot) and metafor v2.4-0 (Egger test, meta-regression). Data and code for all analyses are available in the Supplement.

## RESULTS

### Quality and bias of the studies included

Based on the search terms, 5417 studies were imported for screening. Of those, 2196 were found to be duplicates and were removed. Of the remaining, 3029 were omitted because of misalignment with the primary outcome (e.g. studies of lifestyle interventions to improve quality of life, not weight). Reasons for excluding 177 of the final 184 studies is shown in Figure 1. All authors participated in this process with proportionate agreement ranging from 88-98% and random probability agreement 81-98% (Cohen’s kappa 0.39-0.55). Quality assessment and risk of bias of individual studies was assessed in the context of the primary outcome for the following parameters: sequence generation [5/7 high; 2/7 low], allocation concealment [4/7 high; 3/7 low], blinding of participants and personnel [6/7 high; 1/7 low], blinding of outcomes assessment [3/7 high; 4/7 low], incomplete outcome data [3/7 high; 3/7 low; 1/7 unsure], selective reporting, and other sources of bias [1/7 low; 6/7 unsure]).

### Characteristics of the studies included

Only 7 studies met our full inclusion criteria (Table 1). ^21-27^ Most studies were conducted in people with obesity who also had risk factors for poor outcomes (i.e. high-risk race/ethnicity, pre-existing type 2 diabetes, prediabetes, hypertension, dyslipidemia or the metabolic syndrome). Five of the 7 studies were conducted in the U.S. – with 6 of 7 being multi-centered - in people 40-60 years old with BMI’s mostly in the 30’s. Two of the studies randomized physicians to receive training in weight management the use of which became the intervention for patients. Three studies randomized patients to anti-obesity medication vs. placebo, whereas two used anti-obesity medication in combination with intensive lifestyle counseling and meal replacements. Two studies reported on more than one active interventions: high vs. low dose of medication ^24^ and basic vs. enhanced lifestyle coaching. ^27^ No study randomized patients to lifestyle modification alone in a setting where the providers had not received additional training on the topic. Control conditions frequently used their respective national or local guidelines to advise patients on healthy eating, physical activity and behavior modification. Time spent on giving this advice was minimal (5-7 minutes) and was intended to resemble “usual care”. Three of the studies ^21,23,25^ reported on the studied intervention at only one time-point, while the remaining studies reported on weight loss at different time points. Two of the studies ^23,26^ reported outcome information on completers (Completer Analysis) and we could not ascertain the intention-to-treat status of the outcome in one study. ^22^ The remaining four studies reported outcomes using modified intention-to-treat.

**Table 1.** Studies included in this meta-analysis.

| Citation | Unique inclusion criteria | Site(s) | Age | BMI | control | Intervention | Duration | Weight change |
| --- | --- | --- | --- | --- | --- | --- | --- | --- |
| Boesten<br>2012 | T2D or IFG | 29 sites in the Netherlands | 59.2 | 32.2 | NHG Dutch guidelines (no strict diet; patient sets goals and is monitored) | Rimonabant 20 mg qd | 12 months | +0.18 kg control vs. - 4 kg in intervention |
| Cohen<br>1991 | hypertension | 1 site in the U.S. | 59.5 | 34.1 | “usual care” | Physicians trained to counsel patients on low calorie food choices | 6 & 12 months | +0.56 kg control vs. - 1.8 kg in intervention at 6 mo; +1.33 kg control vs. - 0.88 kg in intervention at 12 mo |
| Davis<br>Martin<br>2006 | Low income African American women | 2 sites in the U.S. | 42 | 39 | Physicians received 2h training on NHLBI guidelines | Physicians received 7h additional training on behavior change for weight loss and multi-disciplinary team made tailored recommendations | 6 months | +0.25 kg control vs. - 1.44 kg in intervention |
| Hauptman<br>2000 |  | 17 sites in the U.S. | 42.4 | 36 | Hypocaloric diet and limit alcohol | Control + orlistat 60 mg or 120 mg QAC | 6 & 12 months | -4.70 kg control vs. - 6.92 kg in low dose and -8.0 kg in high dose intervention at 6 mo; - |
|  |  |  |  |  |  |  |  | 4.14 kg control vs. - 7.08 kg in low dose and -7.94 kg in high dose intervention at 12 mo |
| Lindgarde 2000 | T2D, hypertension or dyslipidemia | 33 sites in Sweden | 53.4 | 33.2 | Mildly hypocaloric diet, 30 min/day walking, placebo pill | Control + Orlistat 120 mg QAC | 12 months | -4.3 kg control vs. - 5.6 kg in intervention |
| Ryan 2010 | $BMI \geq 40 \text{ kg/m}^2$ | 7 sites in the U.S. | 47.1 | 46.1 | Mayo Clinic guidelines for healthy weight | Low calorie liquid diet, then meal replacements, anti-obesity medication and group behavioral therapy | 12 & 24 months | -1.1 kg control vs. - 17.2 kg intervention at 12 mo; - 0.5 kg control vs. - 12.7 kg intervention at 24 mo. |
| Wadden 2011 | Metabolic syndrome | 6 sites in the U.S. | 51.5 | 38.5 | Low calorie diet, 180 min/wk activity, 5-7 min advice from PCP every 3 mo | Control + 10-15 min advice from lifestyle coach every 3 mo, meal replacements and/or anti-obesity medications | 6, 12, 18 & 24 months | -2 kg control vs. - 6.6 kg intervention at 6 mo; - 2.3 kg control vs. - 7.1 kg intervention at 12 mo; - 1.9 kg control vs. - 5.8 kg intervention at 18 mo; - 1.7 kg control vs. - 4.6 kg intervention at 24 mo. |

### Meta-analysis for weight loss outcomes

The seven studies included 2,187 individuals and reported 2,400 changes from baseline in the intervention arm and 2,453 in the control arm. Forest plot of the seven studies is shown in Figure 2. There was evidence of substantial heterogeneity (I^2^ was 86%, τ^2^ was 7.19 and p< 0.01). The fixed effect model estimated a treatment effect for the aggregate efficacy of primary care interventions −2.82 kg (95% CI: −3.27 kg to −2.38 kg), that was substantially different from the random effect estimate of −3.54 kg (95% CI: −5.61 kg to −1.47 kg). Considerable uncertainty remains about the efficacy of interventions; the 95% prediction interval for the treatment effect in future replications of such trials was rather wide: from nearly 11 kg of weight loss to nearly 4 kg of weight gain.

**Figure 2:**
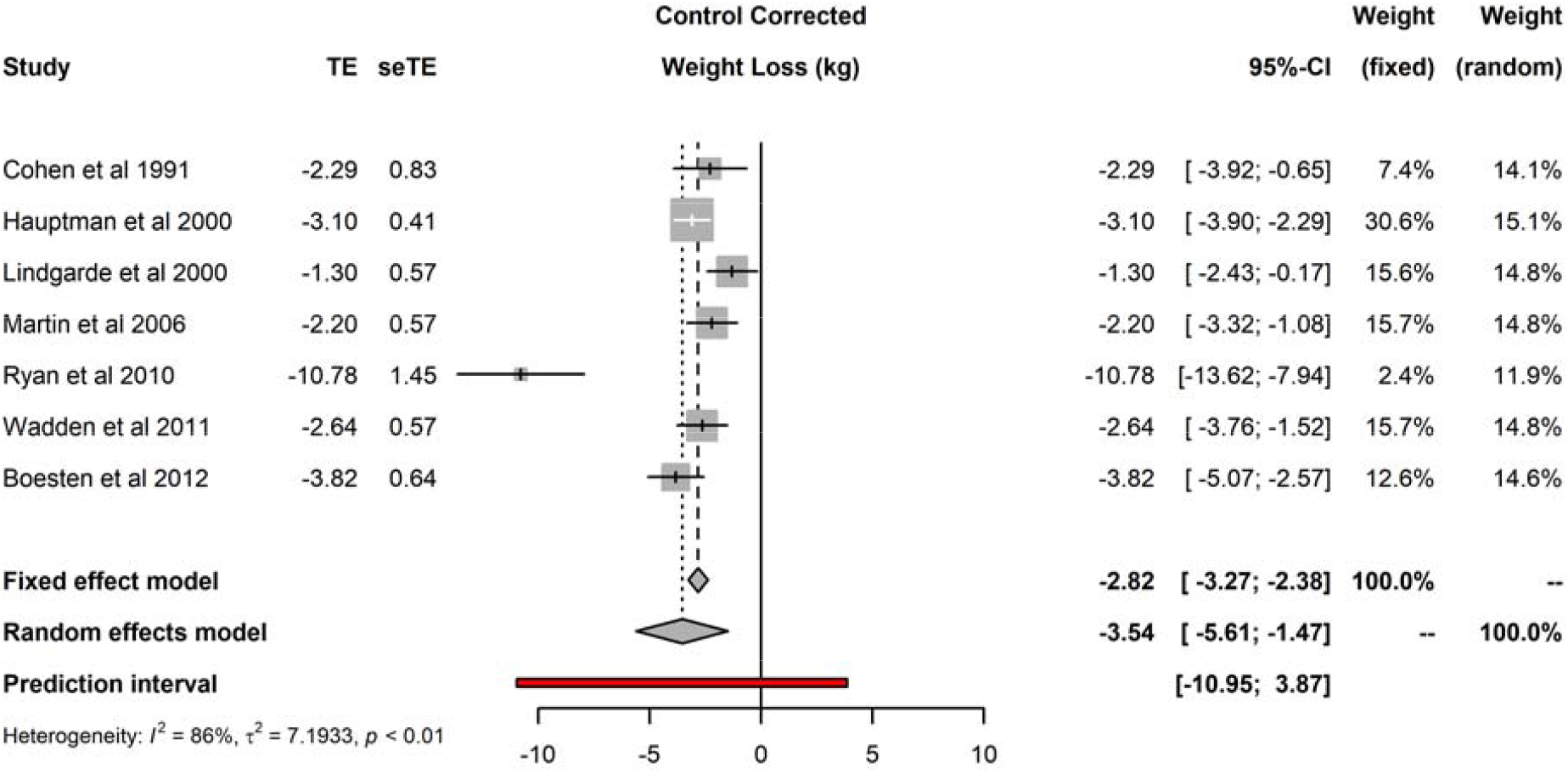
Forest plot of control corrected change in body weight from baseline

Funnel plot analyses (Figure 3) suggested the presence of publication bias: the plot itself was asymmetric, and there appeared to be missing studies in the area of non-significant studies (white area of the plot). In confirmation of these visual impressions, Egger’s test was also statistically significant (p=0.0005). Summary treatment effects for the studies reporting multiple interventions at multiple time points is shown in Figure 4. For two of the trials reported, there was evidence of treatment heterogeneity within each study. ^26,27^

**Figure 3:**
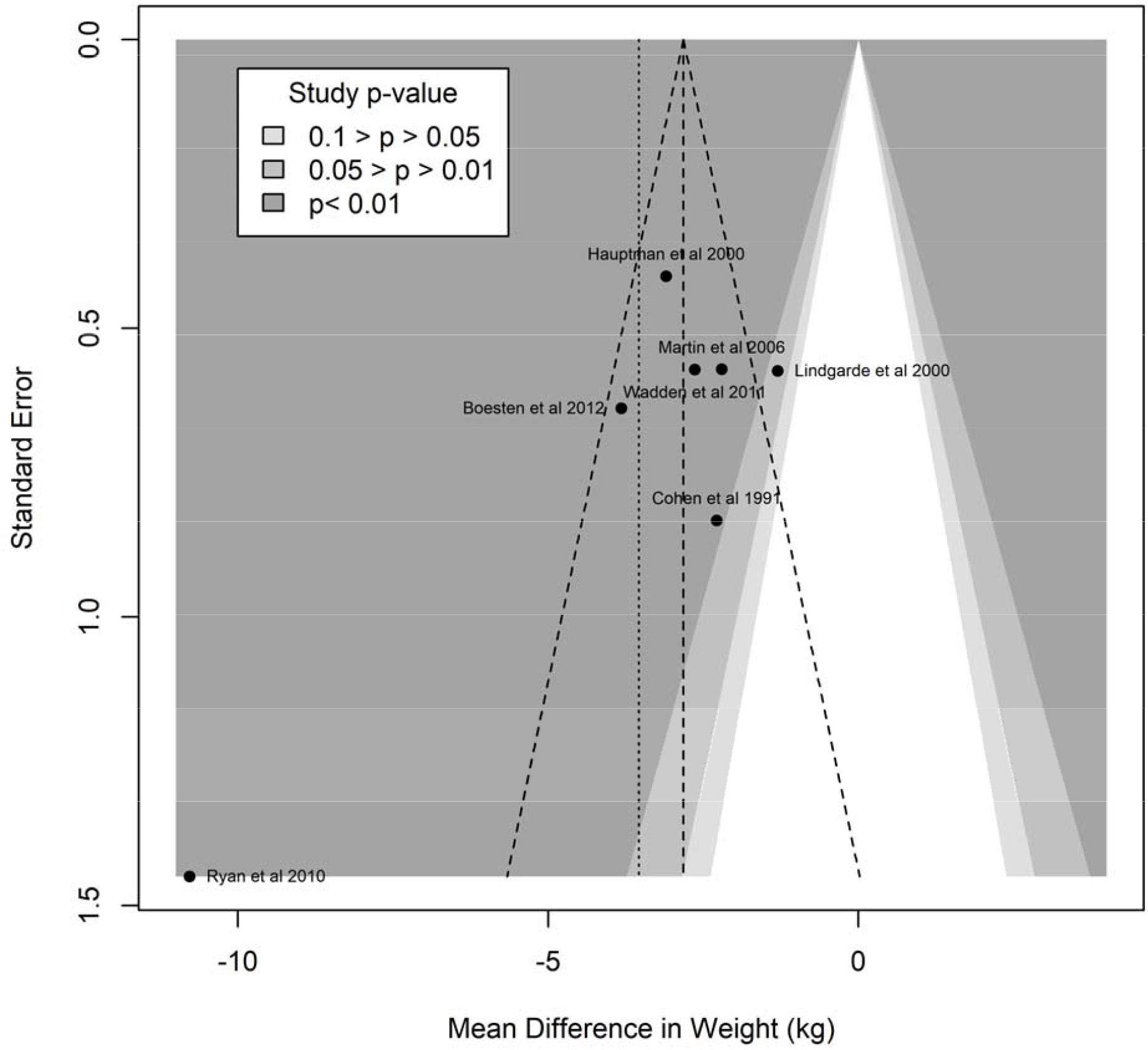
Enhanced funnel plot for assessment of study heterogeneity

**Figure 4.**
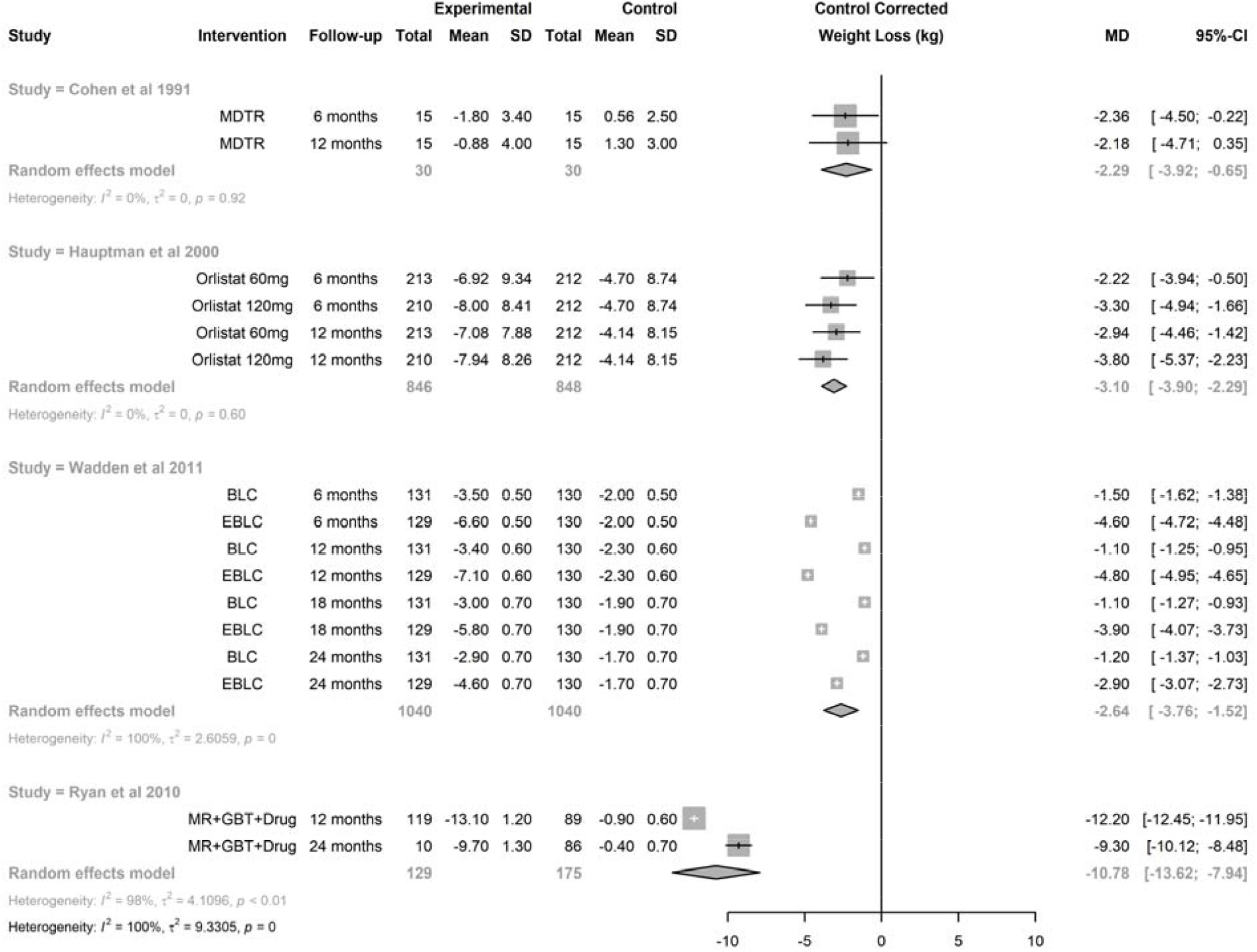
Synthesis of treatment effect from studies reporting multiple interventions at multiple time points BLC: Brief Lifestyle Coaching; EBLC: Enhanced Basic Lifestyle Coaching; GBT: Group Behavioral Therapy; MDTR: Physician Training; MR: Meal Replacement

### Analyses of heterogeneity

In univariate multilevel modeling, using random effects at the study and follow-up (nested within study levels), there was evidence for attenuation of the treatment effect with time, i.e. a reduction in weight loss of about 0.5 kg per six months (p=0.04). Interventions that included a medication component (either alone or as components of multipronged intervention) achieved a greater weight reduction by −2.94 kg (p<0.0001). In multivariate multilevel meta-regressions that included both covariates (type of intervention and duration of the intervention), the estimated weight loss associated with a drug component was identical to that estimated by the univariate model. In these models, there was also evidence for reduction of efficacy with time (reduction in weight loss by 0.53 kg per six months, 95%CI: 0.04-1.0 kg, p-value=0.04). When we analyzed intervention type by treatment time interactions, there was evidence for differential attenuation of treatment effect for interventions that included a drug component. While the loss of efficacy of interventions without a drug component was estimated to be 0.26 kg/6 months (95% CI −0.14 to 0.66 kg, p=0.20), the efficacy of interventions with drug components attenuated at a rate of 0.44 kg/6months, 95%CI (0.34-0.53 kg/6 months, p-value <0.001).

## DISCUSSION

Obesity – with its many comorbid conditions – has now surpassed smoking as the leading cause of preventable death in the United States.^28^ Despite the fact that obesity is both treatable and preventable, treating the comorbidities, rather than obesity *per se* has been the mainstay of therapy. Reasons for lack of weight management prioritization are extensive and complex but have included lack of clinician education on effective obesity management and processes that systematically address weight loss and weight loss maintenance long-term.^6-8^ There are reasons to believe, however, that the paradigm may be shifting. Clinical trials testing infrastructure for weight-prioritized visits in primary care (e.g. NCT04678752) coupled with highly effective pharmacotherapy ^29,30^ may make patient weight loss of 15-22% achievable. Major findings from the current analysis show that patients lose ∼3 kg (approximately 3% body weight) when they are provided more than usual care by their primary care provider and that the amount of weight loss approximates 6 kg (roughly 6% body weight) when anti-obesity medication is part of the intervention. Nevertheless, weight regain is common underscoring the need for long-term treatment and intensification.

Most people receive their health care in primary care – not by specialists, dieticians, or health coaches – the vast majority of whom fail to maintain a normal body weight. ^1^ Although some published data supports the use of commercial weight loss programs ^31^, their long-term use is uncommon. Patients want help with their weight from their primary care provider and 77% of those who receive help (vs. 33% that did not) ^32^ report positive behavior change and weight loss. ^33^ The current analysis examined what *has been (hence could be) done* in primary care for weight management. Interventions ranged from provider use of newly reinforced knowledge in the area of behavior modification to multi-faceted approaches that included intensive behavioral therapy, anti-obesity medications and meal replacements. Wide breadth in the interventions tested led to substantial and residual heterogeneity in their treatment effect. The random effects model estimated a mean patient weight loss of −3.54 kg. Interventions that included a medication component (either alone or as components of multipronged intervention) achieved a greater weight reduction by −2.94 kg. The latter finding was unchanged when adjusting for all components of the intervention or its duration supporting the independent benefit of anti-obesity medications. Together, multi-faceted approaches appear to yield greater weight loss compared to lifestyle advice alone when administered by a primary care provider, however, larger trials are needed to confirm the magnitude of the benefit.

Weight nadir in clinical trials is consistently achieved between 6-12 months with weight regain following, despite ongoing intensive lifestyle modification ^34,35^ or pharmacotherapy. ^36,37^ Results from the current analysis are highly aligned with those previously reported. We observed an average weight regain of 0.5 kg/6 months in the setting of ongoing active intervention. Interestingly, despite greater weight loss being achieved in trials testing anti-obesity medication, the weight regain was also greater (0.44 kg/6 months vs. 0.26 kg/6 months). The notion that rapid (vs. gradual) weight loss leads to greater weight regain has been debunked ^38,39^. Hence, the greater weight regain with anti-obesity medication use may relate to the greater absolute weight loss achieved. Highly conserved evolutionary adaptations for survival are sensitive to weight loss, including lowering energy expenditure ^40^ and driving energy intake. ^41^ Hence, discontinuation of anti-obesity medication after weight loss could explain the rapid weight regain. Advances in the field have made it clear that successful treatment must be long-term.

This systematic review and meta-analysis ultimately revealed few studies quantifying patient weight loss resulting from an intervention administered by a primary care provider. Those that have exhibited considerable heterogeneity and carry uncertainty around whether the results can be replicated. The included studies are collectively of moderate quality, but there appears to be a bias toward missing non-significant studies, hence this analysis may over-estimate the true effect of the interventions. An important strength, however, was that this analysis was not limited to trials examining behavior modification for weight loss nor did it include auxiliary personnel providing care. ^42^ It was intended to review the full breadth of what has been (hence could be) done in routine medical practice using conventional workflow and established scope of training for primary care providers.

It is worth noting that the control conditions were intended to simulate “usual care” – citing medical guidelines or giving brief advice – widely acknowledging the lack of standard-of-care for weight management. High intra-individual variability in response to weight loss interventions ^43^ rightfully supports a patient-centered approach. Nevertheless, lack of consensus on recommendations for a specific approach ^44^ perpetuates clinical inertia. Trials included in this analysis that used established guidelines or “usual care” as the standard-of-care rendered <1 kg of weight loss. ^21-23,26^ Trials included in this analysis that utilized hypocaloric diets and targets for physical activity as the standard-of-care rendered 2-5 kg of weight loss ^24,25,27^ making it clear that prescribed behavior modification is essential. Granularity as to the details of the optimal behavior modification, however, remain elusive.

## CONCLUSIONS

It is time to reframe the conversation such that weight management is the primary goal for treating weight-related comorbidities. ^45^ To that end, this is the first systematic review and meta-analysis to see what *has or could be done* by primary care providers for weight management. Despite the highly pragmatic nature of this question, few clinical trials have pursued the answer. Results demonstrate successful patient weight loss, particularly when multiple strategies are used together. Results also show that weight regain is common, especially after using anti-obesity medication – likely because of discontinuation. It is clear that establishment of a standard-of-care is desperately needed as a first step toward treatment. Layering of additional strategies and long-term treatment must follow to promote patient weight loss and weight loss maintenance.

## Data Availability

Data and code are provided as a supplement to the paper.

## Supplement. Data and code for the meta-analysis

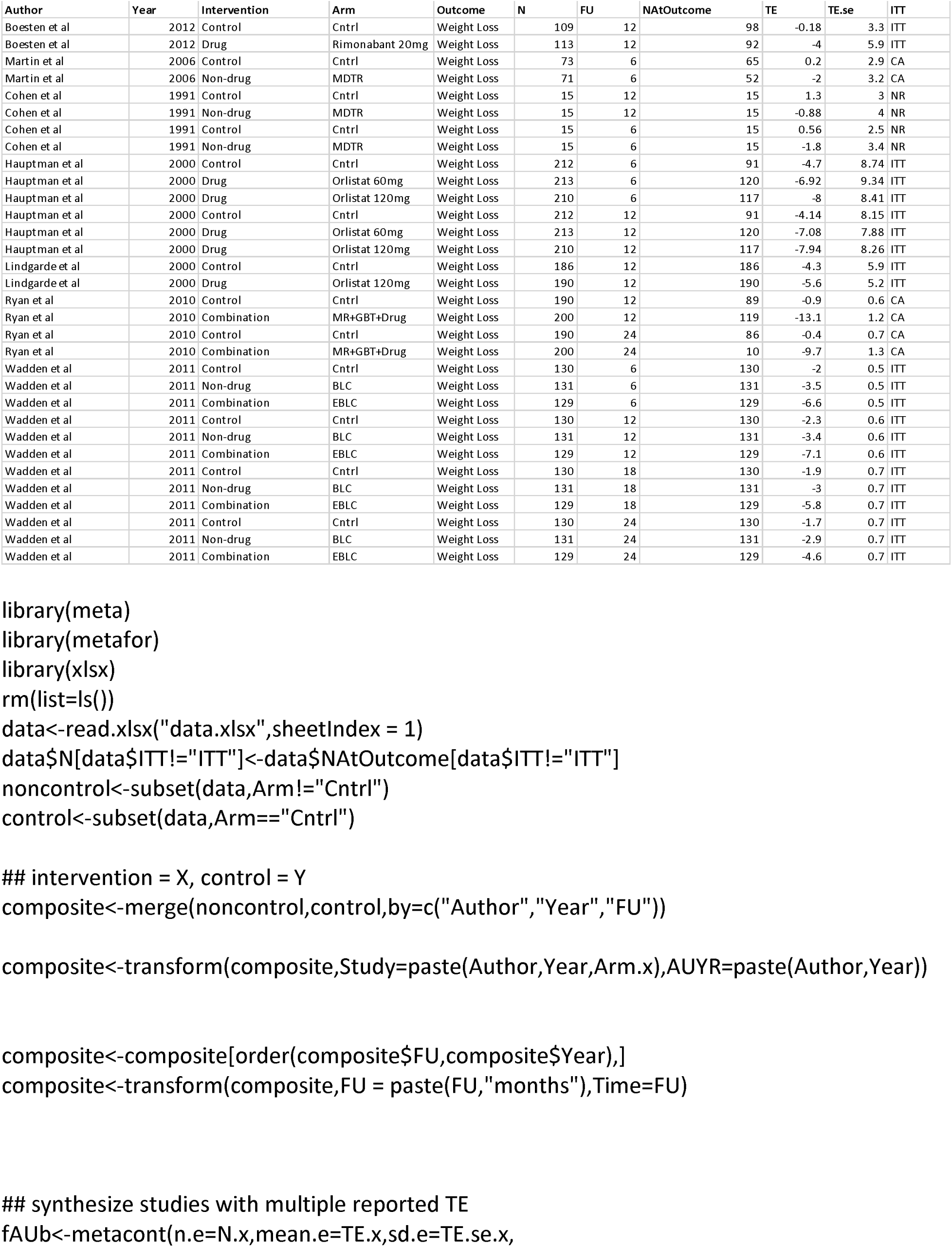
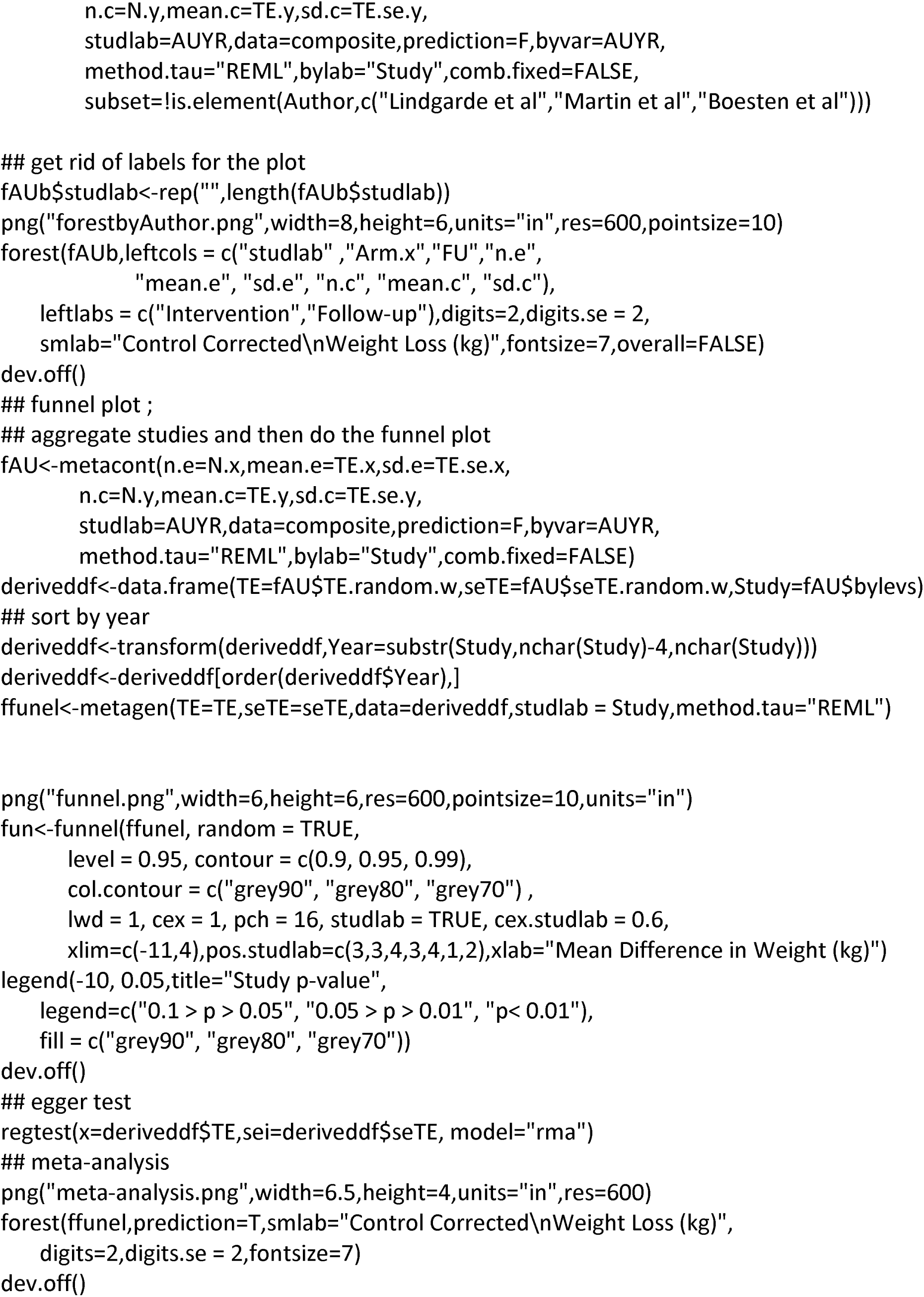
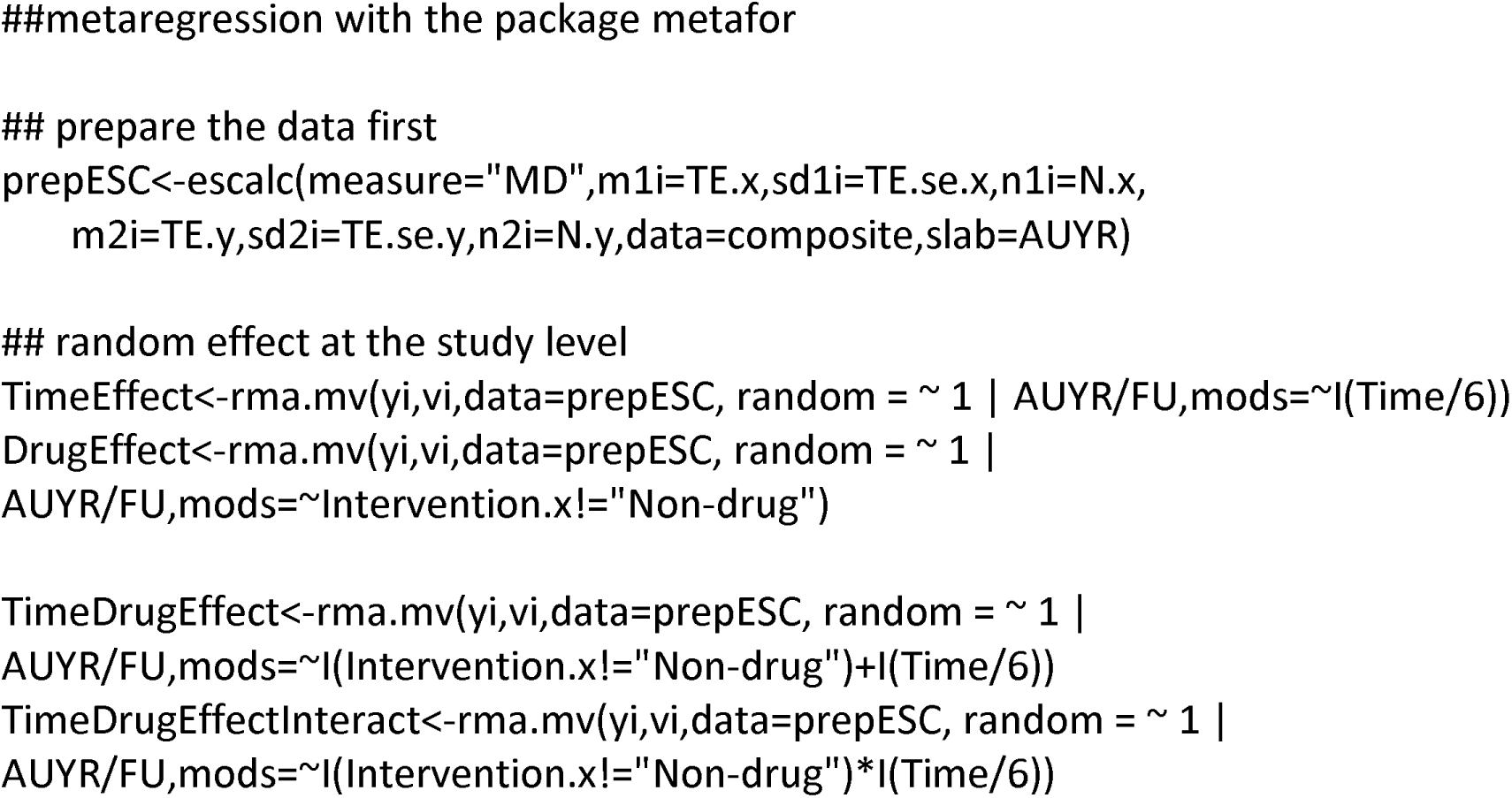

## Notes

### Competing Interest Statement

The authors have declared no competing interest.

### Funding Statement

The study did not receive any funding

### Summary of Updates

Updated discussion

