## Supplementary material for "A closer look at weight loss interventions in primary care: A systematic review and meta-analysis": PROSPERO protocol for meta-analysis (from another version of this preprint: version 1, 2022-08-03)

### Systematic review

This record cannot be edited because it is being assessed by the editorial team

#### 1. * Review title.

Give the title of the review in English

Clinical trials examining the efficacy of weight loss interventions in primary care: A systematic review and meta-analysis

#### 2. Original language title.

For reviews in languages other than English, give the title in the original language. This will be displayed with the English language title.

English

#### 3. * Anticipated or actual start date.

Give the date the systematic review started or is expected to start.

01/02/2021

#### 4. * Anticipated completion date.

Give the date by which the review is expected to be completed.

31/07/2021

#### 5. * Stage of review at time of this submission.

Tick the boxes to show which review tasks have been started and which have been completed.
Update this field each time any amendments are made to a published record.

**Reviews that have started data extraction (at the time of initial submission) are not eligible for inclusion in PROSPERO**.
If there is later evidence that incorrect status and/or completion date has been supplied, the published PROSPERO record will be marked as retracted.

This field uses answers to initial screening questions. It cannot be edited until after registration.

The review has not yet started: No

| Review stage | Started | Completed |
|---|---|---|
| Preliminary searches | Yes | No |
| Piloting of the study selection process | No | No |
| Formal screening of search results against eligibility criteria | No | No |
| Data extraction | No | No |
| Risk of bias (quality) assessment | No | No |
| Data analysis | No | No |

Provide any other relevant information about the stage of the review here.

#### 6. * Named contact.

The named contact is the guarantor for the accuracy of the information in the register record. This may be any member of the review team.

Emileigh Willems

Email salutation (e.g. "Dr Smith" or "Joanne") for correspondence:

Dr. Willems

#### 7. * Named contact email.

Give the electronic email address of the named contact.

#### 8. Named contact address

**PLEASE NOTE this information will be published in the PROSPERO record so please do not enter private information, i.e. personal home address**

Give the full institutional/organisational postal address for the named contact.

13001 East 17th Place, 4th Floor West, Mail Stop B119, Aurora, CO 80045

#### 9. Named contact phone number.

Give the telephone number for the named contact, including international dialling code.

01-806-283-2700

#### 10. * Organisational affiliation of the review.

Full title of the organisational affiliations for this review and website address if available. This field may be completed as 'None' if the review is not affiliated to any organisation.

University of Colorado Anschutz Medical Campus

Organisation web address:

www.ucdenver.edu

#### 11. * Review team members and their organisational affiliations.

Give the personal details and the organisational affiliations of each member of the review team. Affiliation refers to groups or organisations to which review team members belong.

**NOTE: email and country now MUST be entered for each person, unless you are amending a published record.**

Dr Emileigh Willems. Department of Biostatistics and Informatics, Colorado School of Public Health, University of Colorado-Denver Anschutz Medical Campus, Aurora, CO

Dr Leigh Perreault. Department of Medicine; Division of Endocrinology, Metabolism and Diabetes; University of Colorado Anschutz Medical Campus, Aurora, CO USA

Dr Christos Argyropoulos. Department of Medicine; Division of Nephrology; University of New Mexico School of Medicine, Albuquerque, NM USA

Dr Krithika Suresh. Department of Biostatistics and Informatics, Colorado School of Public Health, University of Colorado-Denver Anschutz Medical Campus, Aurora, CO

Dr Peter Smith. Department of Medicine; University of Colorado Anschutz Medical Campus, Aurora, CO USA

##### 12. * Funding sources/sponsors.

Details of the individuals, organizations, groups, companies or other legal entities who have funded or sponsored the review.

The NIH through the funding described below.

Grant number(s)

State the funder, grant or award number and the date of award

NIH funded grant, 1R18DK127003, awarded 09/17/20-07/31/25

##### 13. * Conflicts of interest.

List actual or perceived conflicts of interest (financial or academic).

Yes

Leigh Perreault has received personal fees for consulting and/or speaking from Novo Nordisk, Sanofi, Elli Lilly, Boehringer Ingelheim, Astra Zeneca, Merck, Medscape, WebMD, UpToDate.

##### 14. Collaborators.

Give the name and affiliation of any individuals or organisations who are working on the review but who are not listed as review team members. **NOTE: email and country must be completed for each person, unless you are amending a published record.**

##### 15. * Review question.

State the review question(s) clearly and precisely. It may be appropriate to break very broad questions down into a series of related more specific questions. Questions may be framed or refined using PI(E)COS or similar where relevant.

What is the efficacy of behavior/lifestyle and/or drug interventions for weight loss in overweight and obese adult patients in the primary care setting?

##### 16. * Searches.

State the sources that will be searched (e.g. Medline). Give the search dates, and any restrictions (e.g. language or publication date). Do NOT enter the full search strategy (it may be provided as a link or attachment below.)

English manuscripts in MEDLINE, EMBASE, Web of Science, Cochrane Central from inception to February 28, 2021

##### 17. URL to search strategy.

Upload a file with your search strategy, or an example of a search strategy for a specific database, (including the keywords) in pdf or word format. In doing so you are consenting to the file being made publicly accessible.

Or provide a URL or link to the strategy. Do NOT provide links to your search **results**.

https://www.crd.york.ac.uk/PROSPEROFILES/242344_STRATEGY_20210312.pdf

Do not make this file publicly available until the review is complete

##### 18. * Condition or domain being studied.

Give a short description of the disease, condition or healthcare domain being studied in your systematic review.

Obesity and overweight individuals

##### 19. * Participants/population.

Specify the participants or populations being studied in the review. The preferred format includes details of both inclusion and exclusion criteria.

Inclusion criteria: Patients in the primary care setting with Age >= 18 and BMI >= 25

Exclusion criteria: Patients unable to participate in weight loss treatment due to factors such as cognitive deficits, non-home residence, or limited life expectancy

#### 20. * Intervention(s), exposure(s).

Give full and clear descriptions or definitions of the interventions or the exposures to be reviewed. The preferred format includes details of both inclusion and exclusion criteria.

We will include studies looking at behavior based (exercise, eating habits) and/or medication based weight loss interventions.

#### 21. * Comparator(s)/control.

Where relevant, give details of the alternatives against which the intervention/exposure will be compared (e.g. another intervention or a non-exposed control group). The preferred format includes details of both inclusion and exclusion criteria.

The controls for behavior based studies would be the "usual advice" given by general practitioners (also called standard of care), or would be the "placebo" group for drug intervention studies.

#### 22. * Types of study to be included.

Give details of the study designs (e.g. RCT) that are eligible for inclusion in the review. The preferred format includes both inclusion and exclusion criteria. If there are no restrictions on the types of study, this should be stated.

Randomized clinical trials (RCTs) and any previous meta-analyses of RCTs will be considered for inclusion.
We will not include studies that use some other disease status as an enrollment criterion (e.g., only individuals with Type 2 Diabetes were invited to participate in the study)

#### 23. Context.

Give summary details of the setting or other relevant characteristics, which help define the inclusion or exclusion criteria.

Clinical trials assessing weight loss interventions in overweight/obese individuals that are conducted in a primary care setting.

#### 24. * Main outcome(s).

Give the pre-specified main (most important) outcomes of the review, including details of how the outcome is defined and measured and when these measurement are made, if these are part of the review inclusion criteria.

Patient weight (or weight loss) after trial intervention

* Measures of effect

effect size estimate and its standard error (likely from a linear model), adjustments will be made when necessary to ensure these estimates represent/estimate the same thing (we will need to account for duration of the trial intervention)

#### 25. * Additional outcome(s).

List the pre-specified additional outcomes of the review, with a similar level of detail to that required for main outcomes. Where there are no additional outcomes please state ‘None’ or ‘Not applicable’ as appropriate to the review

None

* Measures of effect

None

#### 26. * Data extraction (selection and coding).

Describe how studies will be selected for inclusion. State what data will be extracted or obtained. State how this will be done and recorded.

The software Covidence will be used for all steps relevant to this question. For abstract screening, two reviewers will independently screen all abstracts for inclusion/exclusion and will be blinded to each other’s decisions. Any disagreements between these two reviewers will be resolved by a third individual/reviewer. The full-text screening process will follow these same steps.

The following elements will be pulled during the data extraction phase using Covidence: study design (e.g., RCT), statistical methodology used (e.g., regression modeling), type of weight loss intervention(s) implemented, covariates accounted for in statistical model or randomization scheme, sample size (total and for each treatment group), demographic information summaries (sex, race/ethnicity, baseline weight/BMI, age), effect size and standard error estimates for primary outcomes (patient weight and/or weight loss after 6, 12 months).

If this information is unavailable/missing or we need additional information to ensure consistency of the data gathered from each study, we will first look at the supplemental/appendix materials of the manuscript and then contact the authors for more information if needed. If the data needed to include the study in the meta-analysis cannot be obtained, the study will be excluded from the meta-analysis, and we will document all steps and reasoning for this decision in the manuscript.

One individual will extract the data using Covidence, with another individual checking their work. A third individual will resolve any disagreements, and a biostatistician will be very involved in this phase to ensure the correct and needed data is acquired. The software Covidence will be used for the data extraction process, and the data can be easily exported from Covidence to the format needed by the biostatistician for the subsequent meta-analysis.

#### 27. * Risk of bias (quality) assessment.

State which characteristics of the studies will be assessed and/or any formal risk of bias/quality assessment tools that will be used.

We will use the Cochrane Risk of Bias tool for RCTs through Covidence. Two review authors will independently assess the risk of bias of each study by evaluating: (1) Adequacy of the randomization scheme/strategy, (2) completeness and consistency of the outcome data (e.g., problematic attrition, exclusion criteria, clear confounders, etc.), (3) possible or clear selective outcome reporting, and (4) adequate blinding of participants and/or clinicians when appropriate. The design of each study will be closely evaluated for any other potential biases by our two reviewers (biostatistician and endocrinologist). A third reviewer will resolve any disagreements that cannot be settled between the two reviewers.

Bias will also be assessed at the outcome level, where possible issues for our outcome (weight loss) could include measurement inconsistencies between individuals or inadequate statistical modeling (clear model assumption violations, unaccounted for covariates/confounders, needed transformations, etc.). If the bias of a study is found to be too extreme, it will be excluded. A small bias issue could disqualify a study from the meta-analysis, but not be severe enough to remove the study entirely from the systematic review. In either case, all details leading to and justifying the decision will be given.

#### 28. * Strategy for data synthesis.

Describe the methods you plan to use to synthesise data. This **must not be generic text** but should be **specific to your review** and describe how the proposed approach will be applied to your data.
If meta-analysis is planned, describe the models to be used, methods to explore statistical heterogeneity, and software package to be used.

Our biostatistician, who has a background in meta-analysis methods, will lead and implement all analyses described in this section. Since we are gathering information for all weight loss interventions implemented in primary care, we expect to have results from numerous behavior-based and drug-based interventions. To adequately model these differences and evaluate all possible indirect comparisons available, we will implement a network meta-analysis using the weight loss effect size and standard error estimates for each study.

Both local and global approaches will be used to evaluate the incoherence/inconsistency of the studies/interventions, where the SIDE (Separating Indirect from Direct Evidence) approach will be implemented local evaluation and the Q-test and $I^2$ statistic will be referenced for global evaluation. All of these inconsistency measures will be implemented using the netmeta package in R. Sub-networks will be considered if the level of inconsistency is found to be too large in the full network (e.g., sub-network of drug-based interventions only, sub-network of behavior-based interventions only).

If the level of inconsistency and number of studies for each intervention is found to be inadequate for any type of network meta-analysis, we will instead perform random effects meta-analyses separately by intervention using the R package meta. Comparisons between the interventions would then be evaluated using prediction intervals calculated for each intervention.

#### 29. * Analysis of subgroups or subsets.

State any planned investigation of ‘subgroups’. Be clear and specific about which type of study or participant will be included in each group or covariate investigated. State the planned analytic approach.

To ensure that the effect size and standard error estimates from each study are estimating the same value and modeled on the same scale, we will evaluate if patients’ age, sex, race/ethnicity, and baseline BMI were adequately modeled in each study included in our meta-analysis. Similarly, the effect size estimates will be corrected for any differences in trial/intervention duration across studies. Once the effect size estimates are all on the same scale, differences in intervention type will be modeled using a network meta-analysis (details described in the previous question).

#### 30. * Type and method of review.

Select the type of review, review method and health area from the lists below.

Type of review

| | |
|---|---|
| Cost effectiveness | No |
| Diagnostic | No |
| Epidemiologic | No |
| Individual patient data (IPD) meta-analysis | No |
| Intervention | No |
| Meta-analysis | Yes |
| Methodology | No |
| Narrative synthesis | No |
| Network meta-analysis | No |
| Pre-clinical | No |
| Prevention | No |
| Prognostic | No |
| Prospective meta-analysis (PMA) | No |
| Review of reviews | No |
| Service delivery | No |
| Synthesis of qualitative studies | No |
| Systematic review | Yes |
| Other | No |

Health area of the review

| | |
|---|---|
| Alcohol/substance misuse/abuse | No |
| Blood and immune system | No |

| | |
|---|---|
| Cancer | No |
| Cardiovascular | No |
| Care of the elderly | No |
| Child health | No |
| Complementary therapies | No |
| COVID-19 | No |
| Crime and justice | No |
| Dental | No |
| Digestive system | No |
| Ear, nose and throat | No |
| Education | No |
| Endocrine and metabolic disorders | Yes |
| Eye disorders | No |
| General interest | No |
| Genetics | No |
| Health inequalities/health equity | No |
| Infections and infestations | No |
| International development | No |
| Mental health and behavioural conditions | No |
| Musculoskeletal | No |
| Neurological | No |
| Nursing | No |
| Obstetrics and gynaecology | No |
| Oral health | No |
| Palliative care | No |
| Perioperative care | No |
| Physiotherapy | No |

| | |
|---|---|
| Pregnancy and childbirth | No |
| Public health (including social determinants of health) | No |
| Rehabilitation | No |
| Respiratory disorders | No |
| Service delivery | No |
| Skin disorders | No |
| Social care | No |
| Surgery | No |
| Tropical Medicine | No |
| Urological | No |
| Wounds, injuries and accidents | No |
| Violence and abuse | No |

#### 31. Language.

Select each language individually to add it to the list below, use the bin icon to remove any added in error.

English

There is an English language summary.

#### 32. * Country.

Select the country in which the review is being carried out. For multi-national collaborations select all the countries involved.

United States of America

#### 33. Other registration details.

Name any other organisation where the systematic review title or protocol is registered (e.g. Campbell, or The Joanna Briggs Institute) together with any unique identification number assigned by them.
If extracted data will be stored and made available through a repository such as the Systematic Review Data Repository (SRDR), details and a link should be included here. If none, leave blank.

#### 34. Reference and/or URL for published protocol.

If the protocol for this review is published provide details (authors, title and journal details, preferably in Vancouver format)

No I do not make this file publicly available until the review is complete

#### 35. Dissemination plans.

Do you intend to publish the review on completion?

Yes

We will submit the final mauscript for consideration to the Journal of the American Medical Association.

#### 36. Keywords.

Give words or phrases that best describe the review. Separate keywords with a semicolon or new line. Keywords help PROSPERO users find your review (keywords do not appear in the public record but are included in searches). Be as specific and precise as possible. Avoid acronyms and abbreviations unless these are in wide use.

Overweight
obesity
weight management
primary care

#### 37. Details of any existing review of the same topic by the same authors.

If you are registering an update of an existing review give details of the earlier versions and include a full bibliographic reference, if available.

#### 38. * Current review status.

Update review status when the review is completed and when it is published.
New registrations must be ongoing so this field is not editable for initial submission.

Review_Ongoing

#### 39. Any additional information.

Provide any other information relevant to the registration of this review.

#### 40. Details of final report/publication(s) or preprints if available.

Leave empty until publication details are available OR you have a link to a preprint (NOTE: this field is not editable for initial submission).
List authors, title and journal details preferably in Vancouver format.
